# Underestimation of Blood Pressure and Stroke Risk by Manual Blood Pressure Measurement

**DOI:** 10.64898/2026.02.24.26346929

**Authors:** Carolina Lopez-Silva, Aditya Surapaneni, Jung-Im Shin, Leora I. Horwitz, Saul B. Blecker, Carina M. Flaherty, Kathryn Foti, Morgan E. Grams, Alexander R. Chang

## Abstract

**Background:** Hypertension guidelines recommend the use of automated BP devices over manual devices to reduce observer bias, such as terminal digit preference. We aimed to evaluate systematic differences in BP readings and the association with incident stroke according to type of measurement.

**Methods:** Using de-identified electronic health record data from Optum Labs Data Warehouse from primary care visits in 2024, we classified providers’ BP measurement method using proportion of odd terminal digit preference as a proxy for manual devices (defined as <0.5% odd digits) and automated devices (defined as 45-55% odd digits). Patients from the manual and automated groups were matched on demographic and clinical covariates. We evaluated cross-sectional BP distributions by measurement modality, and compared mean BP and proportions meeting clinical thresholds using t-tests, and chi squared tests, respectively. In a separate 2019 cohort created using the same methods, we evaluated whether longitudinal associations between systolic BP and incident stroke differ by measurement method.

**Results:** Among 336,634 matched patients, mean SBP in the automated group was 131.7 (19.3) mmHg and 125.9 (14.8) mmHg in the manual group. The absolute percentage of patients meeting BP clinical thresholds differed substantially (<130/80: automated 33.2% vs. manual 38.8%; <140/90: automated 61.2% vs. manual 70.9%). Among 686,482 matched patients in the 2019 cohort, the manual group had a 1.16-fold (1.10-1.22) higher risk of stroke at any given BP compared to the automated group.

**Conclusion:** Manual BP measurement was associated with lower mean BP, BP control, and potential underestimation of stroke risk.

## INTRODUCTION

High blood pressure is one of the most prevalent modifiable risk factors for cardiovascular disease. In the United States, an estimated 47.7% of adults age 18 and older, and 71.6% of adults 60 and older, meet criteria for hypertension (≥130 mmHg systolic blood pressure or ≥80 mmHg diastolic blood pressure or use of antihypertensive medication).(1) Hypertension is associated with a higher risk of stroke, coronary artery disease, heart failure, atrial fibrillation, and all-cause mortality.(2) Accordingly, greater efforts have been made toward earlier diagnosis of hypertension and more rigorous blood pressure control, as reflected in the 2025 multi-society hypertension guidelines.(3) Paramount to timely detection and treatment of hypertension is the accurate measurement of blood pressure, with the guidelines recommending the use of automated oscillometric blood pressure devices when measuring in-office blood pressure in adults. (3)

There are several reasons that professional societies favor automated office blood pressure measurements over the traditional auscultatory method. First, mercury was phased out due to health and environmental concerns, and thus auscultatory methods rely on aneroid devices that require frequent calibration (every 6 months for wall-mounted; every 2-4 weeks for mobile devices).(4) In addition, multiple studies have documented evidence of human error or observer bias with the auscultatory method, such as terminal digit preference, in which readings are rounded to end on “0”.(5,6) Terminal digit preference itself has been associated with lower recorded blood pressure values and a corresponding reduction in the number of appropriate hypertension diagnoses.(7–9) This effect can ultimately result in a pattern of underdiagnosis and subsequent undertreatment of hypertension, particularly in settings where manual auscultatory techniques remain the primary mode of measurement.

In this study, we aimed to evaluate systematic differences in blood pressure readings and the association with adverse outcomes according to type of blood pressure measurement. Because the specific device type is not typically recorded in administrative or clinical datasets, we used terminal digit preference for even numbers as a proxy for use of manual measurement, Manual aneroid sphygmomanometers require visual estimation of Korotkoff sounds, and the World Health Organization (WHO) recommends rounding to the nearest even number when using these devices, leading to the vast majority of blood pressure values ending in even digits(10). A prior study by Kottke et al. demonstrated that this pattern clearly exists for manual devices and resolves when clinics transition from manual devices to automated devices, supporting terminal digit distribution as an indicator of measurement modality(7). We also compared mean blood pressure values between patients seen by providers using manual measurements and those using automated measurements. Lastly, we assessed whether the association between blood pressure and incident stroke differed between the two measurement modalities, choosing stroke as the adverse outcome most strongly associated with elevated blood pressure(11).

## METHODS

### Study Population

This study used de-identified administrative claims and electronic health record (EHR) data from the Optum Labs Data Warehouse (OLDW). The database contains longitudinal health information on enrollees and patients, representing a mixture of ages and geographical regions across the United States. The claims data in OLDW includes medical and pharmacy claims, laboratory results and enrollment records for over 350M commercial and Medicare Advantage enrollees. The EHR-derived data includes a subset of EHR data that has been normalized and standardized into a single database(12). We included patients with a blood pressure reading at a primary care site (outpatient visits in family medicine, general practice, internal medicine or primary care as defined in Optum) between January 1^st^, 2024 through December 31^st^, 2024. We further limited our sample to patients who received care from a provider who we could classify as likely to have exclusively used a manual method or an automated method for BP measurement. We randomly selected one primary care visit per person. To classify providers, we required recorded blood pressure measurements on at least ten patients during the study period and quantified the frequency of odd terminal digits. Based on examination of distribution of terminal digits and consistent with prior findings on the distribution of terminal digits in manual versus automated blood pressure readings(7), providers with <0.5% odd digits across patients were classified as manual device users, and providers with 45-55% odd digits across patients were classified as automated device users **(Supplemental Figure 1)**.

To evaluate the association between blood pressure readings and incident stroke, a separate cohort of patients with primary care visits in 2019 were selected using the same approach and followed through December 31^st^, 2024. The primary analyses used the 2024 dataset to leverage the most recent data, while the 2019 dataset was used to define the stroke incidence cohort to ensure sufficient longitudinal follow-up for outcome ascertainment.

### Covariates

We ascertained hypertension and diabetes history based on relevant International Classification of Disease, 9^th^ and 10^th^ Modification (ICD-9 or ICD-10) diagnosis codes, with ≥1 inpatient code, ≥1 problem-list code, or ≥2 outpatient codes within a 2-year period (**Supplemental Table 1).** Demographic characteristics, insurance category, smoking status, body mass index (BMI), and use of antihypertensive medications at baseline were also determined.

### Blood Pressure Measurements and Outcomes

Blood pressure measurements were abstracted from the electronic health record. If more than one blood pressure was recorded at a given visit, the first measurement was selected for analysis. Incident stroke was determined as abstracted ICD-9 and ICD-10 codes from the inpatient record (ICD-9: 431.xx, 432.xx, 433.xx, 434.xx; ICD-10: I61.xx, I62.xx, I63.xx). As a negative control outcome, we also included incident hospitalization with vehicular accidents (ICD9: E810–E829; ICD10: V00–V89).

### Statistical Methods

Characteristics of the study population were described using mean +/- standard deviation (SD) for continuous variables and frequencies and percentages for categorical variables. To ensure comparability of patients receiving manual vs. automated blood pressure measurements, propensity scores for manual measurement were determined using logistic regression and age, sex, race, insurance type (including a category for missing insurance status), history of diabetes, history of hypertension, BMI, smoking history, and use of antihypertensive medication as covariates; patients from the manual and automated group then underwent 1:1 nearest neighbor propensity score matching without replacement, using a caliper of 0.0005. Patients missing BMI (<2% of the total population) were not included in the matched sample. This process was performed separately for both the 2024 and 2019 cohorts.

Blood pressure measurement values were compared between the manual and automated groups using t-tests, as well as within categories of blood pressure control using chi-squared distributions. In the matched 2019 cohort, Cox proportional hazards regression was used to evaluate the association of SBP with incident stroke, with a reference value of 120mmHg measured with an automated device were used. Patients were followed from the time of blood pressure measurement to first stroke, death, last clinical encounter, or December 31, 2024, whichever occurred first. Blood pressure was modeled as a cubic spline with 5 knots. To assess for potential residual confounding, we also assessed the association of measurement modality with a negative control outcome of incident vehicular accidents using the same approach. Finally, because manual blood pressure readings may preferentially exclude the initial measurement in routine practice, we conducted a sensitivity analysis evaluating automated and manual blood pressure using the second recorded measurement among people who had more than one measurement during an encounter.

## RESULTS

### Baseline Characteristics of Study Population

Among 8,661 providers with at least 10 blood pressure measurements in 2024, 2,500 were classified as using manual or automated devices as described above with the remainder likely using a mix of automated and manual devices to record blood pressure. A total of 776,209 patients had an encounter in 2024 with a provider classified as strictly using manual or automated blood pressure devices **(Supplemental Figure 2)**. Baseline characteristics are summarized in **Table 1**. In the 468,632 patients with automated measurements, the mean (SD) age was 57.2 (17.4) years, 59.9% were women, and 15.3% were Black. In the 307,677 with manual measurements, the mean (SD) age was 58.9 (17.1) years, 56.3% were women, and 9.3% were Black.

**Table 1.** Baseline characteristics of patient population. A portion of individuals underwent propensity score matching based on age, sex, race, insurance type, history of diabetes, history of hypertension, BMI, smoking history, and use of antihypertensive medication. Data presented as mean (standard deviation) and frequency (%). Baseline characteristics were compared between manual and automated groups using t-tests for continuous variables and chi-squared distributions for categories. SMD = standardized mean difference.

| Characteristic | Unmatched |  |  | Propensity Score Matched |  |  |
| --- | --- | --- | --- | --- | --- | --- |
|  | Automated | Manual | SMD | Automated | Manual | SMD |
| *N* | 468632 | 307577 |  | 168317 | 168317 |  |
| Age | 57.2 (17.4) | 58.9 (17.1) | 9% | 59.5 (16.6) | 58.9 (17.3) | -4% |
| Women | 280556 (59.9%) | 173218 (56.3%) | -7% | 95501 (56.7%) | 95435 (56.7%) | 0% |
| White | 300585 (64.1%) | 255975 (83.2%) | 42% | 142227 (84.5%) | 144171 (85.7%) | 3% |
| Black | 71496 (15.3%) | 28465 (9.3%) | -18% | 16494 (9.8%) | 14967 (8.9%) | -3% |
| Hispanic | 49302 (10.5%) | 7889 (2.6%) | -30% | 3314 (2.0%) | 3027 (1.8%) | -1% |
| Asian | 22113 (4.7%) | 7355 (2.4%) | -12% | 3354 (2.0%) | 3408 (2.0%) | 0% |
| Other Race | 25136 (5.4%) | 7893 (2.6%) | -14% | 2928 (1.7%) | 2744 (1.6%) | -1% |
| Commercial Insurance | 210555 (44.9%) | 145861 (47.4%) | 5% | 88203 (52.4%) | 90397 (53.7%) | 3% |
| Medicaid | 39727 (8.5%) | 16157 (5.3%) | -12% | 8844 (5.3%) | 9527 (5.7%) | 2% |
| Medicare | 133550 (28.5%) | 74947 (24.4%) | -9% | 45909 (27.3%) | 45029 (26.8%) | -1% |
| Missing Insurance | 70536 (15.1%) | 64868 (21.1%) | 16% | 22020 (13.1%) | 19422 (11.5%) | -5% |
| Other Insurance | 5207 (1.1%) | 3120 (1.0%) | -1% | 1981 (1.2%) | 2109 (1.3%) | 1% |
| Uninsured | 9057 (1.9%) | 2624 (0.9%) | -9% | 1360 (0.8%) | 1833 (1.1%) | 3% |
| SBP (mmHg) | 129.6 (18.8) | 125.6 (14.6) | -23% | 131.7 (19.3) | 125.9 (14.8) | -33% |
| DBP (mmHg) | 77.6 (10.3) | 76.3 (9.4) | -13% | 78.2 (10.2) | 76.5 (9.4) | -17% |
| Antihypertensive Use | 194211 (41.4%) | 149512 (48.6%) | 14% | 86527 (51.4%) | 85889 (51.0%) | -1% |
| Hypertension | 254519 (54.3%) | 174401 (56.7%) | 5% | 99007 (58.8%) | 98369 (58.4%) | -1% |
| Diabetes | 110661 (23.6%) | 70759 (23.0%) | -1% | 39689 (23.6%) | 40224 (23.9%) | 1% |
| Current Smoker | 65541 (14.0%) | 52104 (16.9%) | 8% | 27533 (16.4%) | 28035 (16.7%) | 1% |
| Former Smoker | 115785 (24.7%) | 73103 (23.8%) | -2% | 43168 (25.6%) | 43430 (25.8%) | 0% |
| BMI (kg/m2)* | 29.9 (7.2) | 30.7 (7.4) | 11% | 30.6 (7.5) | 30.6 (7.4) | 1% |

\*BMI was missing in <2% of the participants.

### Comparison of Blood Pressure Measurements Derived from a Manual or Automated Method

Of the 776,209 patients in the original cohort, 336,634 were successfully matched between the manual and automated groups. Among matched patients, the mean measured SBP in the automated group was 131.7 (19.3) mmHg, compared to 125.9 (14.8) mmHg in the manual group; corresponding values for diastolic blood pressure were 78.2 (10.2) in the automated group and 76.5 (9.4) in the manual group **(Figure 1)**. The percentage of patients with SBP below clinical thresholds also differed between groups: <120 mmHg (27.3% in automated vs. 30.3% in manual), <130 mmHg (49.4% in automated vs. 62.3% in manual) and <140 mmHg (70.8% in automated vs. 83.7% in manual) (all p<0.001). Similarly, the percentage of patients meeting BP goals incorporating systolic and diastolic BP differed: (BP <130/80 mmHg 33.2% in automated vs. 38.8% in manual; BP <140/90 mmHg 61.2% in automated vs. 70.9% in manual). Across automated blood pressure measurements, the distribution of terminal digits was evenly spread across 0 to 9 (9-11% for each digit), while in the manual group, 31.2% of the readings ended in 0 **(Figure 2).** Repeat blood pressure measurements were recorded more often in the automated group (57.6% vs. 22.3%, p <0.001), with similar distributions in the second measurement compared to the first measurement. In sensitivity analyses using the second blood pressure measurement for those with more than one measurement at a visit, mean systolic blood pressure remained higher with automated compared to manual measurement (129.6 vs 125.5 mmHg), consistent with the primary analysis. Results were consistent among unmatched patients (data not shown).

**Figure 1.**
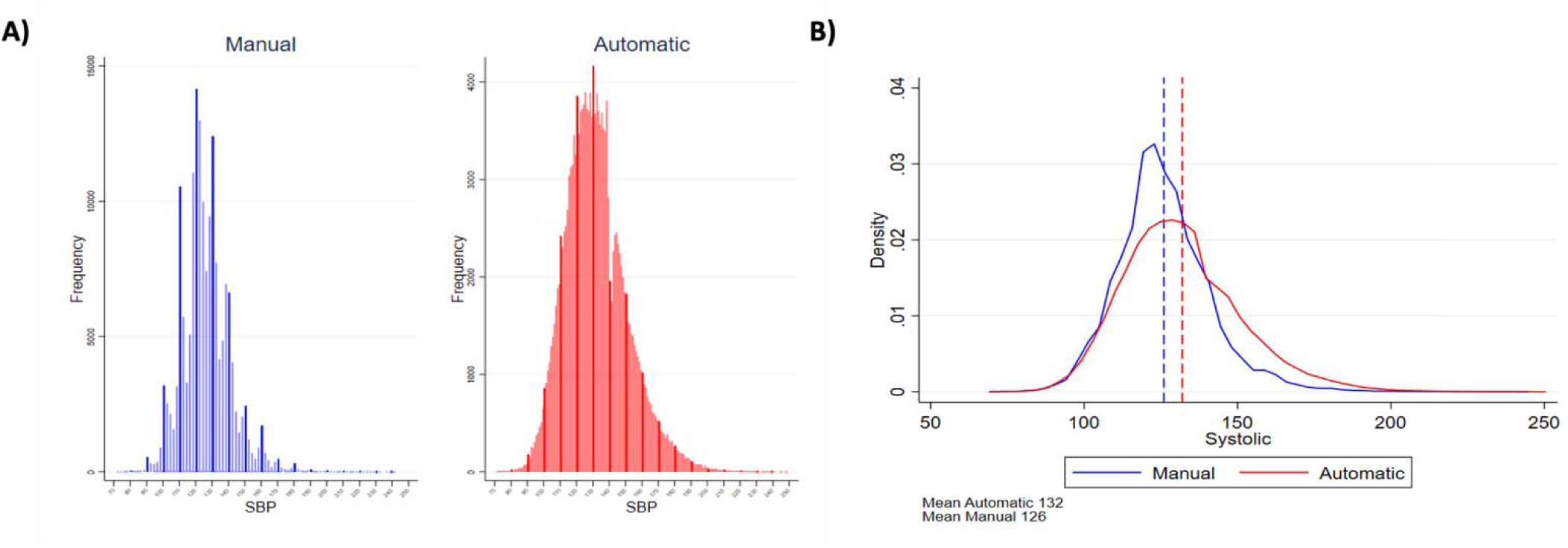
**A)** Histograms (bars with accentuated color used for measurements ending in 0) and **B)** density plot of distribution of systolic blood pressure (mmHg) measurements in the manual and automated groups.

**Figure 2.**
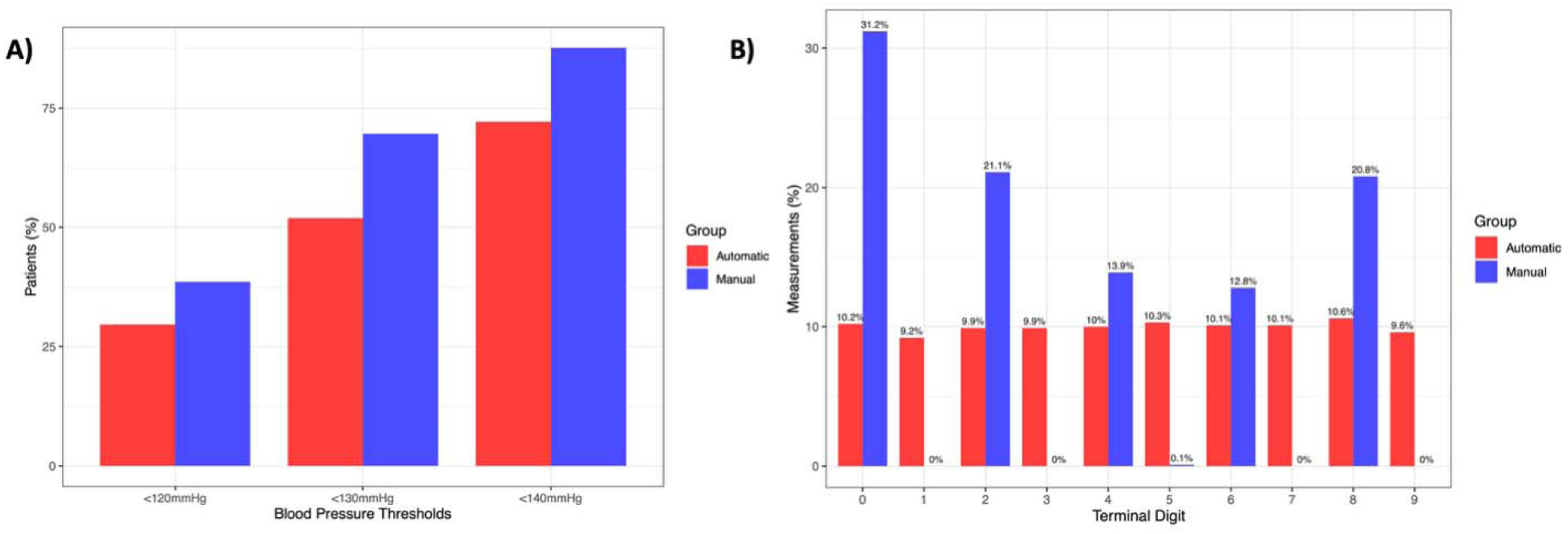
**A)** Bar graphs of percent of patients with systolic blood pressures below clinically relevant thresholds (120mmHg, 130mmHg, 140mmHg) in automatic and manual groups. **B)** Distribution of terminal digit in manual and automated groups.

### Association of Blood Pressure Measured with a Manual or Automated Method with Incident Stroke

The baseline characteristics of the 1,912,473 patients with a primary care visit in 2019 to a provider classified as strictly manual or strictly automated are summarized in **Supplemental Table 2.** After propensity matching, 686,482 remained for subsequent analyses, with 5,107 incident strokes occurring over a mean follow-up time of 5.7 years. Consistent with the initial cohort, mean SBP was 131.3mmHg (19.1) in the automatic group and 124.6mmHg (15.6) in the manual group; mean DBP was 77.6mmHg (10.8) in the automatic group and 75.8mmHg (10.0) in the manual group. For both groups, at values above 110mmg, higher SBP was associated with a higher risk of incident stroke (**Figure 3**). For a given level of stroke risk, the corresponding blood pressure was lower when measured manually than when measured automatically. For example, compared to a reference SBP of 120mmHg measured automatically, an automatic SBP of 125 had a HR 1.16 [1.10 - 1.22], while a manual SBP of 120 had a HR of 1.16 [1.09 – 1.22] **(Figure 3).** In terms of the negative control outcome of incident vehicular accidents, there was no significant difference in accidents in the matched automatic (1.20%) and manual (1.19%) groups (HR 1.01 [95% CI: 0.97 - 1.06], p=0.60).

**Figure 3.**
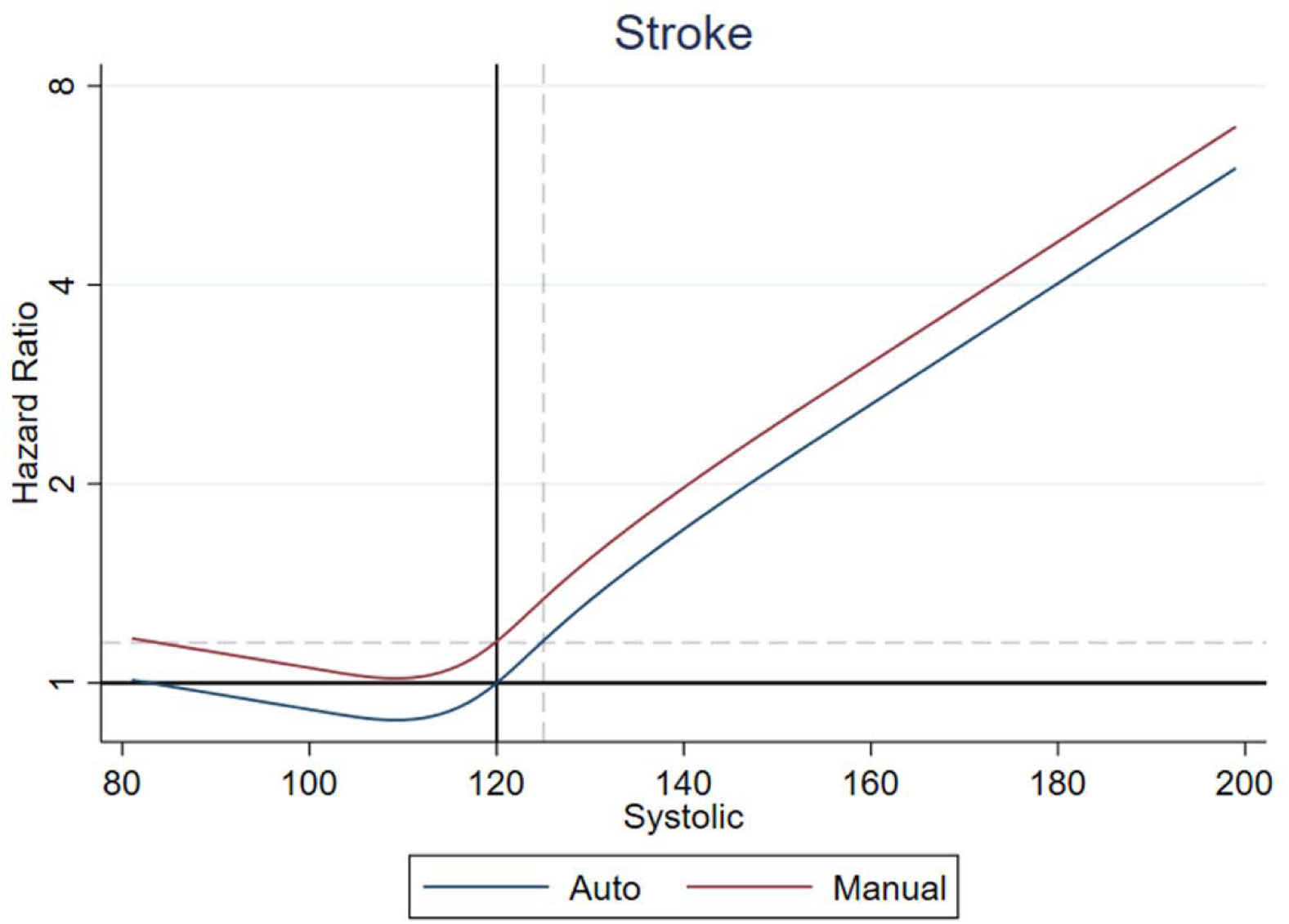
Association of systolic blood pressure (mmHg) and hazard ratio of incident stroke among patients with automated and manual blood pressure measurements. Gray dash line illustrates stroke risk for an automated blood pressure of 125mmHg is comparable to a manual blood pressure of 120mmHg.

## DISCUSSION

This study highlights the potential impact of blood pressure measurement modality in achieving optimal blood pressure control and reducing cardiovascular risk in primary care practices. Measuring blood pressure manually was associated with 5.8 mmHg lower mean systolic blood pressure than measuring blood pressure automatically, despite otherwise similar patients. However, for any given blood pressure level, manually measured blood pressures were associated with higher risk of incident stroke than blood pressure readings obtained through automated measurement methods – approximately a 5 mmHg difference. These systematic differences in blood pressure and associated stroke risk between the two groups likely represent human error and observer bias in the manual measurement modality, tending toward a more optimistic assessment of blood pressure control and thus underestimation of stroke risk.

Given the widely prevalent use of manual measurement modalities across the US and globally, these findings have significant implications for public health. While no exact estimate of BP modality use exist, a 2021 study administered to 3 cardiovascular or hypertension professional societies found that 39% of respondents measured BP manually(13), and a 2017 national survey of Canadian family physicians found that only 54% reported using manual office BP measurement(14). Many primary care providers continue to rely on and prefer manual BP measurements, with prior research noting that this preference is often attributed to their training in using this modality(9,15). As shown in our study and in quality improvement studies implementing automated devices, BP measured with automated devices in real-world settings is often associated with higher BP than when measured manually(7,9).

One important finding in our study, consistent with others, is that manual blood pressure readings had an increased rate of digit preference – specifically, for numbers ending in zero – compared to automated methods. Similarly, prior studies found that in clinics using manual BP measurement, 32.8% of readings ended in zero(7). Digit preference has been associated with a lower mean systolic blood pressure and with fewer patients meeting diagnostic criteria for hypertension or receiving pharmacological therapy(7–9). Systematic underestimation of blood pressure thus may promote clinical inertia, preventing appropriate titration of antihypertensive medications, and missing opportunities to prevent downstream consequences of hypertension, including stroke, cardiovascular disease, heart failure, and cognitive decline(16,17). Indeed, a study by Nietert et al. found that patients receiving care at primary care practices with increased rates of terminal digit levels had lower odds (odds ratio 0.92, 95% CI: 0.85, 0.99) of receiving antihypertensive medication prescriptions than patients in practices receiving care at practices with lower terminal digit preference levels(8).

Our study had notable strengths, including its large sample size, the use of propensity matching to minimize confounding, and use of a negative control outcome. In addition, the use of longitudinal data enabled us to assess stroke risk over time rather than relying on cross-sectional associations. On the other hand, we were limited by inability to determine with certainty whether readings were taken manually or in an automated manner; instead, provider practices were inferred based on frequency of blood pressure values ending in an even or odd number. In addition, we were unable to assess other aspects of appropriate blood pressure measurement technique such as waiting for 5 minutes while the patient is not conversing, seated with their feet flat on the floor, back supported, arm at heart level, and using an appropriately sized blood pressure cuff. Finally, a limitation of our analysis is that death is a competing risk; accordingly, our estimates should not be interpreted as true stroke cumulative incidence but rather as risk estimates used to compare manual- and automated-BP–based predictions.

As more provider practices adopt guideline-recommended automated methods for blood pressure measurement, our results suggest that we may initially see a rise in hypertension diagnoses as the influence of human bias is reduced. Of note, few patients had repeat blood pressure measurements, although this occurred more frequently within the automated blood pressure group. Guidelines recommend taking multiple measurements. Improved adoption of optimal blood pressure measurement techniques will enable clinicians to more accurately diagnose and treat hypertension to optimal targets. In turn, improved diagnostic precision may prompt earlier initiation or intensification of therapy, ultimately enhancing cardiovascular outcomes.

## Data Availability Statement

Under agreement with the OLDW we cannot share individual data with third parties.

