## Supplement for "Underestimation of Blood Pressure and Stroke Risk by Manual Blood Pressure Measurement"

**SUPPLEMENTAL MATERIALS**

| **Covariate/Outcome** | **Diagnosis Codes (ICD9/ICD10)** |
| --- | --- |
| Diabetes Mellitus | ICD-9: 250.xx, ICD-10: E10.xx, E11.xx, E13.xx |
| Hypertension | ICD9: 401.xx, ICD10: I10.xx, SNOMED: 40.xx |
| Stroke (Ischemic/Cerebrovascular) | ICD-9: 431.xx, 432.xx, 433.xx, 434.xx, ICD-10: I61.xx, I62.xx, I63.xx |
| Vehicular Accident | ICD9: E810–E829; ICD10: V00–V89 |

**Supplemental Table 1.** Diagnosis codes (ICD9/ICD10) for history of diabetes, hypertension and outcomes of stroke and vehicular accidents.

|  | **Unmatched** | | | | **Propensity Score Matched** | | | |
| --- | --- | --- | --- | --- | --- | --- | --- | --- |
| **Characteristic** | **Automated** | **Manual** | **SMD** | ***P*** | **Automated** | **Manual** | **SMD** | ***P*** |
| *N* | 809713 | 1102760 |  |  | 343241 | 343241 |  |  |
| Age | 54.3 (16.9) | 55.4 (16.9) | 7% | <0.001 | 55.6 (16.9) | 55.7 (17.2) | 0% | 0.402 |
| Women | 476850 (58.9%) | 635801 (57.7%) | -3% | <0.001 | 202484 (59.0%) | 202210 (58.9%) | 0% | 0.501 |
| White | 549394 (67.9%) | 930775 (84.4%) | 40% | <0.001 | 263270 (76.7%) | 256978 (74.9%) | -4% | <0.001 |
| Black | 127189 (15.7%) | 69435 (6.3%) | -31% | <0.001 | 35929 (10.5%) | 38894 (11.3%) | 3% | <0.001 |
| Hispanic | 61723 (7.6%) | 43766 (4.0%) | -16% | <0.001 | 19617 (5.7%) | 22029 (6.4%) | 3% | <0.001 |
| Asian | 29474 (3.6%) | 25360 (2.3%) | -8% | <0.001 | 10595 (3.1%) | 11490 (3.3%) | 1% | <0.001 |
| Other Race | 41933 (5.2%) | 33424 (3.0%) | -11% | <0.001 | 13830 (4.0%) | 13850 (4.0%) | 0% | 0.902 |
| Commercial Insurance | 427004 (52.7%) | 562742 (51.0%) | -3% | <0.001 | 179981 (52.4%) | 175775 (51.2%) | -2% | <0.001 |
| Medicaid | 66887 (8.3%) | 57964 (5.3%) | -12% | <0.001 | 24410 (7.1%) | 25489 (7.4%) | 1% | <0.001 |
| Medicare | 207371 (25.6%) | 258799 (23.5%) | -5% | <0.001 | 91660 (26.7%) | 97297 (28.3%) | 4% | <0.001 |
| Missing Insurance | 61370 (7.6%) | 195512 (17.7%) | 30% | <0.001 | 35694 (10.4%) | 31709 (9.2%) | -4% | <0.001 |
| Other Insurance | 19893 (2.5%) | 7761 (0.7%) | -15% | <0.001 | 3780 (1.1%) | 4057 (1.2%) | 1% | 0.002 |
| Uninsured | 27188 (3.4%) | 19982 (1.8%) | -10% | <0.001 | 7716 (2.2%) | 8914 (2.6%) | 2% | <0.001 |
| SBP (mmHg) | 130.1 (18.8) | 124.5 (15.4) | -33% | <0.001 | 131.3 (19.1) | 124.6 (15.6) | -38% | <0.001 |
| DBH (mmHg) | 77.9 (10.8) | 75.9 (10.0) | -19% | <0.001 | 77.6 (10.8) | 75.8 (10.0) | -17% | <0.001 |
| Antihypertensive Use | 294736 (36.4%) | 458811 (41.6%) | 11% | <0.001 | 150655 (43.9%) | 152512 (44.4%) | 1% | <0.001 |
| Hypertension | 348572 (43.0%) | 494459 (44.8%) | 4% | <0.001 | 158705 (46.2%) | 162305 (47.3%) | 2% | <0.001 |
| Diabetes | 143116 (17.7%) | 188504 (17.1%) | -2% | <0.001 | 62810 (18.3%) | 65544 (19.1%) | 2% | <0.001 |
| Current Smoker | 114058 (14.1%) | 153744 (13.9%) | 0% | 0.004 | 47112 (13.7%) | 44934 (13.1%) | -2% | <0.001 |
| Former Smoker | 184577 (22.8%) | 263117 (23.9%) | 3% | <0.001 | 72098 (21.0%) | 80004 (23.3%) | 6% | <0.001 |
| BMI (kg/m2) | 30.1 (7.4) | 30.5 (7.3) | 5% | <0.001 | 30.3 (7.4) | 30.5 (7.4) | 3% | <0.001 |

**Supplemental Table 2.** Baseline characteristics of the 2019 cohort, used for evaluation of association of systolic blood pressure with incident stroke. A portion of individuals underwent propensity score matching based on age, sex, race, insurance type, history of diabetes, history of hypertension, BMI, smoking history, and use of antihypertensive medication. Data presented as mean (standard deviation) and frequency (%). Baseline characteristics were compared between manual and automated groups using t-tests for continuous variables and chi-squared distributions for categories. SMD = standardized mean difference.

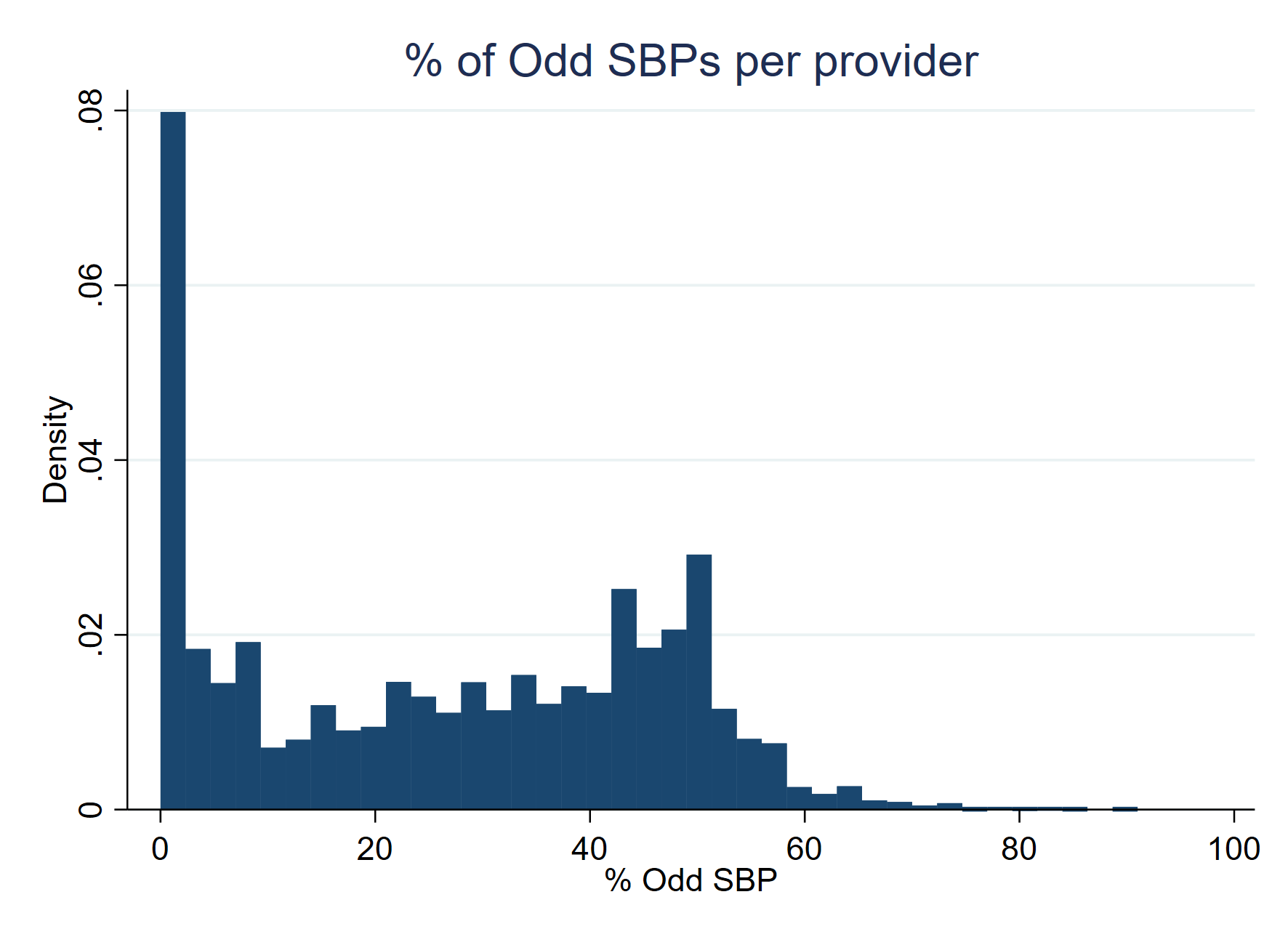

**Supplemental Figure 1.** Percentage of systolic blood pressure (SBP) measurements per provider ending in an odd digit (e.g., 1, 3, 5, 7, 9). Measurements by providers whose practice’s SBP readings ended on an odd number less than 0.5% of the time were defined as ‘manual’, while those from providers whose readings ended in odd digits between 45 to 55% of the time were defined as ‘automated’.

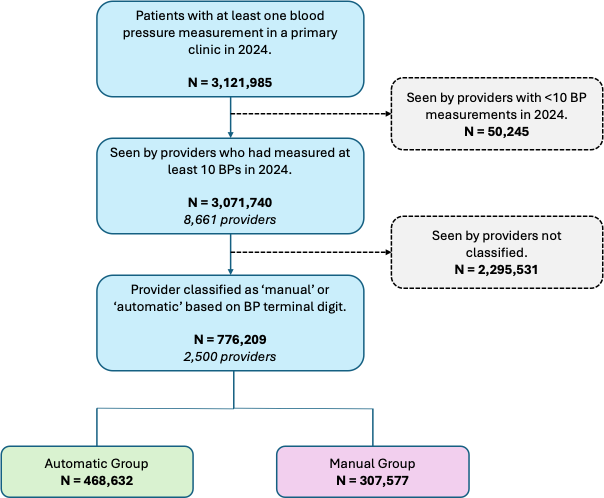

**Supplemental Figure 2.** Population flowchart. Our final study population included 776,209 patients who were seen by providers with at least 10 blood pressure (BP) measurements in 2024 and who were able to be classified as manual or automatic based on the frequency of odd BP terminal digits.
